# Amplitude and Frequency Modulation of EEG Predicts Intraventricular Haemorrhage in Preterm Infants

**DOI:** 10.1101/2024.03.15.24303868

**Authors:** Emad Arasteh, Maria Luisa Tataranno, Maarten De Vos, Xiaowan Wang, Manon J.N.L. Benders, Jeroen Dudink, Thomas Alderliesten

**Author notes:** Correspondence to: Thomas Alderliesten, MD, PhD Wilhelmina Children’s Hospital/University Medical Centre Utrecht, Department of Neonatology, Room KE04.123.1, PO Box 85090,3584 AE Utrecht, The Netherlands. These authors contributed equally to this work (Jeroen Dudink, Thomas Alderliesten).

## Abstract

**Background:** Intraventricular hemorrhage (IVH) is a common and significant complication in premature infants. While cranial ultrasound is the golden standard for IVH detection, it may not identify lesions until hours or days after occurring, which limits early intervention. Predicting IVH in premature infants would be highly advantageous. Recent studies have shown that EEG data’s amplitude and frequency modulation features could offer predictive insights for neurological diseases in adults.

**Methods:** To investigate the association between IVH and EEG monitoring, a retrospective case-control study was conducted in preterm infants. All infants underwent amplitude integrated EEG monitoring for at least 3 days after birth. The study included 20 cases who had an IVH diagnosed on cranial ultrasound and had a negative ultrasound 24h earlier, and 20 matched controls without IVH. Amplitude and frequency modulation features were extracted from single-channel EEG data, and various machine learning algorithms were evaluated to create a predictive model.

**Results:** Cases had an average gestational age and birth weight of 26.4 weeks and 965 grams, respectively. The best-performing algorithm was adaptive boosting. EEG data from 24 hours before IVH detection proved predictive with an area under the receiver operating characteristic curve of 93%, an accuracy of 91%, and a Kappa value of 0.85. The most informative features were the slow varying instantaneous frequency and amplitude in the Delta frequency band.

**Conclusion:** Amplitude and frequency modulation features obtained from single-channel EEG signals in extremely preterm infants show promise for predicting IVH occurrence within 24 hours before detection on cranial ultrasound.

## Introduction

The developing brains of preterm infants are extremely vulnerable, which causes preterm infants to have a high risk of long-term neurological sequelae. Prediction and early detection of potential permanent brain injury is critical to ensure timely and appropriate treatment [1]. Intraventricular haemorrhage (IVH) is one of the most serious complications of preterm birth [2]. IVH is commonly graded from 1 to 3, based on the severity of the haemorrhage [3]. An IVH can lead to a venous infarction (previously known as grade 4 IVH) and post-haemorrhagic ventricular dilatation (PHVD), both conditions are linked to neurodevelopmental impairment [4].

Routine cranial ultrasound (cUS) is the gold standard to detect IVH in preterm infants. However, as cUS is intermittent, an IVH might not be detected until hours or even days after the actual occurrence [5, 6]. Timely detection of IVH and its complications is crucial to ensure tailored medical interventions to minimize the risk of (additional) brain injury and thereby improve patient outcomes [5, 7, 8]. Predicting the likelihood of IVH before it occurs is even more valuable than prompt detection. This proactive approach allows for preventive measures to be taken to potentially prevent the onset of IVH altogether.

Recent advances in artificial intelligence (AI) algorithms have enabled more efficient and detailed quantitative analysis of preterm infants’ neural oscillations, providing clinicians with a more efficient and accurate means of identifying infants at risk of neurological sequalae [9, 10]. There is a demand for new innovative feature extraction and learning algorithms to characterize brain signals for prediction, detection, and management of brain injury [11–14]. Automatic analysis of EEG signals by using machine learning and AI techniques have shown great potential for predicting long-term outcomes among neonates [15–17].

Previous studies have demonstrated that EEG recordings can detect alterations in the electrical activity of the brain after developing an IVH. These changes in EEG background have been linked to the severity of IVH [18]. Furthermore, early amplitude-integrated EEG (aEEG) characteristics have been correlated with IVH patterns within the first three days after birth [19], and positive Rolandic sharp waves of premature EEG have been observed in the majority of IVH cases [20]. Nevertheless, despite recent advancements in identifying risk factors associated with IVH, methods that can accurately and dynamically predict IVH in preterm infants before it occurs are lacking [21, 22].

Current predictive and diagnostic approaches for IVH in preterm infants are mainly based on EEG signal burst changes in the time domain, which are more prognostic of outcome than predictive of IVH occurrence. Iyer et al [23] have shown that burst shapes differ significantly for cases with upcoming IVH and infants having no IVH, but they have not provided substantiating results on the length of time windows of prediction before IVH occurrence. Nevertheless, EEG exhibits promising capabilities as a valuable instrument for illustrating a wide range of neuronal activity patterns beyond the burst characteristics commonly linked to brain injury and neurodevelopmental outcomes [24–26]. Therefore, there is a demand for incorporating new brain dynamical features for the prediction of brain damage (like IVH) among preterm infants.

A novel approach is amplitude and frequency modulation (AM-FM) of brain signals, which could provide valuable insights about neural activity and dynamics [27]. The AM-FM processing allows for the detection of nonlinear variations in amplitude, frequency, and power across different frequency bands [28]. Previous studies in adults have demonstrated the ability of AM and FM features to demonstrate differences between healthy and diseased groups for Parkinson’s disease and Alzheimer’s disease [29, 30]. Despite major differences in brain dynamics between neonates and adults, changes in amplitude, frequency, and power across the frequency bands of activity also occur in neonates IVH [31–33].

The primary aim of this study is to utilize AM-FM features of EEG in preterm infants with a gestational age (GA) less than 30 weeks, to predict the occurrence of IVH. By utilizing AM and FM features, we aim to identify new potential biomarkers for prediction of IVH, on EEG data 24 hours before IVH detection. The secondary aims are to identify the most crucial new features for IVH prediction, evaluate the prediction accuracy for each individual subject to provide personalized information for clinical decision-making, and determine the optimal EEG time epoch length for IVH prediction.

## Materials and Methods

### Study Design

A retrospective case-control study was performed on data obtained between 2009 and 2017. The need for written informed consent was waived by the medical ethical board of the University Medical Center (UMC) Utrecht (14-335/C). Cases were matched to controls based on sex, GA, and birthweight. Major risk factors for IVH were compared [34]. Baseline characteristics of IVH cases and matched controls are listed in Table 1.

**Table 1.** Baseline characteristics of 20 IVH cases and 20 matched controls.

|  | **Case** | **Control** |
| --- | --- | --- |
| **Male sex, n (%)** | 15 (75%) | 15 (75%) |
| **Birth weight, g, med (IQR)** | 965 (807 – 1074) | 970 (800 - 1092) |
| **5 min. Apgar score, med (IQR)** | 7 (6 – 8) | 8 (7 - 9) |
| **Gestational age, week, mean (SD)** | 26.4 (1.26) | 26.9 (1.21) |
| **Antenatal CCS, n (%)** |  |  |
| - **Partial course** | 8 (40%) | 7 (35%) |
| - **Complete course** | 12 (60%) | 13 (65%) |
| **Maternal PE/HELLP, n (%)** | 0 (0%) | 2 (10%) |
| **SGA, n (%)** | 1 (5%) | 1 (5%) |
| **HSPDA, n (%)** | 13 (65%) | 7 (35%) |
| **Mech. Ventilation, <72h** | 18 (90%) | 18 (90%) |
| **EOS, n (%)** | 0 (0%) | 0 (0%) |
| **LOS, n (%)** | 8 (42.1%) | 7 (35%) |
| **Surgical NEC, n (%)** | 1 (5%) | 1 (5%) |
| **Surfactant, n (%)** | 19 (95%) | 16 (80%) |
| **Inotropes <72h, n (%)** | 12 (60%) | 8 (40%) |
| **IVH** |  |  |
| - **Grade I/II** | 7 (35%) | - |
| - **Grade III/PVHI** | 13 (65%) | - |

CCS: corticosteroids; EOS: microbiologically proven sepsis <48h of age; IVH: intraventricular hemorrhage; HELLP: hemolysis elevated liver enzymes and low platelets syndrome; HSPDA: hemodynamically significant patent ductus arteriosus; IQR: interquartile range; LOS: microbiologically proven sepsis >48h of age; NEC:

### Cranial Ultrasound for IVH Detection

A cUS was performed on admission and repeated daily. Patients were included when they had no IVH on the cUS at admission but did develop a IVH on subsequent cUS. Patients were only included when there was an IVH negative cUS available the day/24h preceding the IVH positive cUS, while having aEEG data for at least the 24h leading up to IVH detection. The cUS images were reviewed by experienced neonatologists specializing in neonatal neurology and cUS imaging (JD, MLT).

### Amplitude-Integrated EEG

The aEEG, including 1 or 2 channel raw EEG recordings, were performed using the BrainZ BRM 3 monitor (Natus Medical Inc., Seattle, WA). The raw EEG data were acquired at 256 Hz using two electrodes for C3-C4 channel recording and a central electrode as reference. Given that the frontal and parietal lobes are commonly affected by IVH, EEG data obtained from C3-C4 positions is well suited to study in relation to IVH occurrence. Per routine clinical neuromonitoring protocol at the time, all neonates <30 weeks GA were monitored with aEEG until at least 72h after birth.

### Pre-Processing EEG

A bandpass filter of 0–30 Hz was applied. To apply artifact correction (similar to approach in [35]) on the data, each recording was divided into 5-minute epochs. Data points that deviated from the epoch mean by more than 250 μV were discarded and replaced by linear interpolation, unless more than 7000 of such samples (10% of epoch length) were rejected.

### Feature Extraction

Prediction algorithms in neonatology research are highly sensitive to varying EEG epoch lengths [36–38]. Furthermore, sensitivity analysis in small datasets is challenging. To perform sensitivity analysis for epoch length, a 1-h epoch length is hypothesized to work best and validated by boot-strapping the data and computing mutual information. Figure 1 demonstrates the schematic of how AM-FM features are extracted and used for classification.

**Figure 1.**
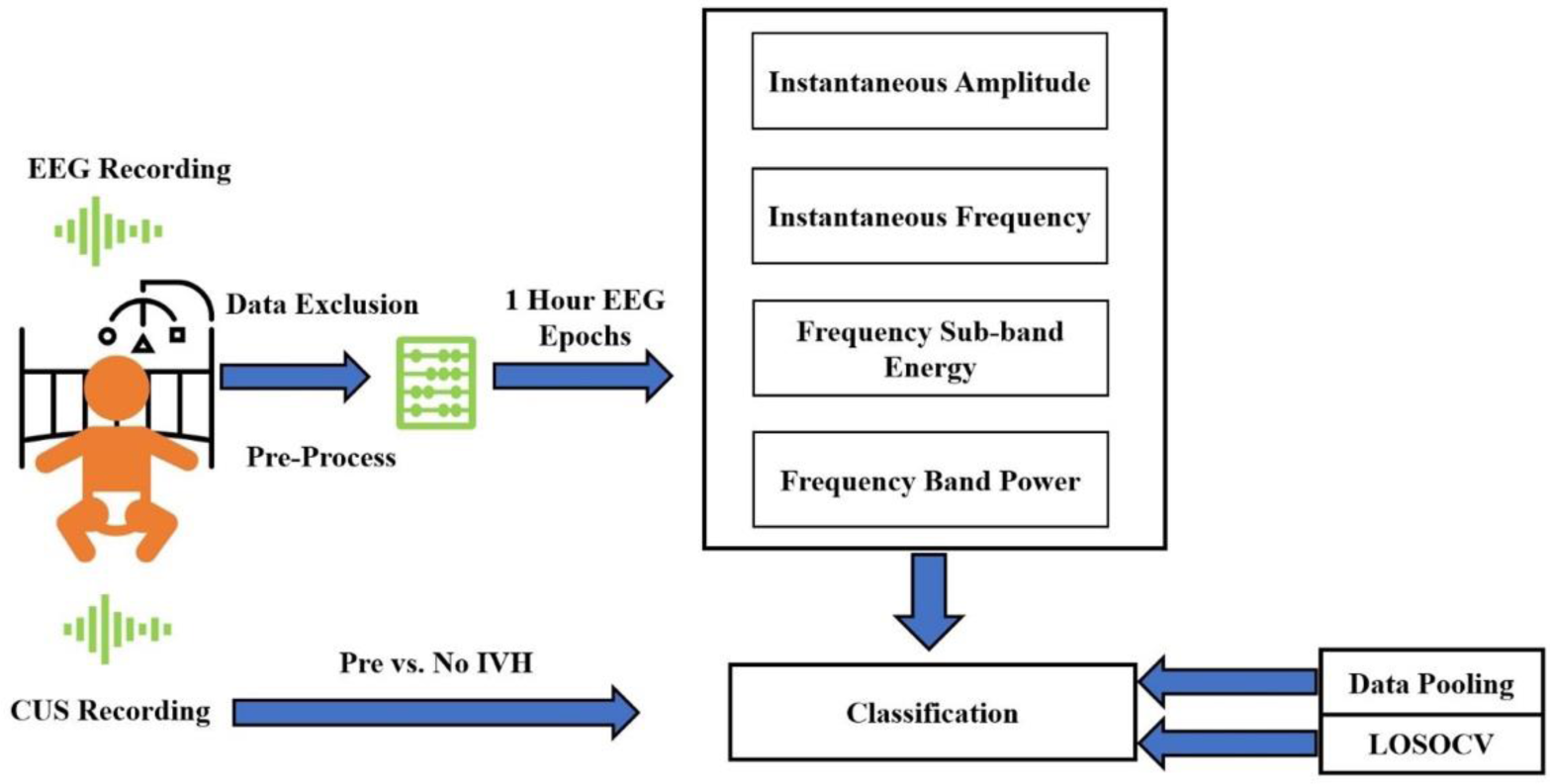
Schematic general overview of IVH and control groups classification steps in this research.

First, AM-FM and AM power features from each 1-h epoch of EEG were extracted. Table 2 shows there are a total of 34 features extracted from each epoch of EEG. For each subject, this yields 24 vectors of features (i.e., 24 1-h epochs of data per subject). Then, these 24*34 (number of EEG epochs*number of features) matrices were pooled for cases and controls (i.e., 40 subjects combined) to apply binary classification. Each of the 34 features was extracted for the same epoch length (e.g., 1-h) based on the equations provided in the supplement. An example of 40 seconds of EEG data and the corresponding AM-FM dynamics of is shown in Supplementary figure S1.

**Table 2.** List of features and their relation to the dynamical state of the brain.

| **Feature** | **Number** | **Description** |
| --- | --- | --- |
| **AM** | 4 (1 for each band, delta, theta, alpha, beta) | Instantaneous amplitude variations |
| **FM** | 4 (1 for each band) | Instantaneous frequency variations |
| **FM slow** | 4 (1 for each band) | Instantaneous slow frequency variations |
| **FM phase slip** | 4 (1 for each band) | Instantaneous fast frequency variations |
| **AM power modulation** | 10 (relative band compared to its domain and previous ones:<br>$\#(4,2) = 10$) | Decoupling fast and slow-varying nature of energy among each frequency band |
| **Relative and absolute power** | 8 (1 for each band) | Power of each band |

The trained model performance was evaluated, and dominant features were determined. The full-band EEG epochs are decomposed into four sub-bands: Delta (0.5-4 Hz), Theta (4-8 Hz), Alpha (8-12 Hz), and Beta (12-30 Hz).

### Classification, Performance, and Parameter Tuning

To differentiate between IVH and the control group, we compared six commonly used machine learning classifiers (using the hyperparameter optimization values described in Supplementary (Table S1)). For a more detailed description of each classifier, refer to Li et al [39]. An Inter- and Intra-Subject analysis were performed by a 10-fold cross-validation, and a leave-one-subject-out cross-validation (LOSOCV) algorithm, respectively. To demonstrate the discriminative power of a single feature towards the output, we also calculated feature importance for the best classifier.

### Inter- and Intra-Subject Analysis

To assess the general validity of the model we conducted a 10-fold cross-validation [40] on the pooled data of 40 subjects. For the 40 subjects, we incorporated 40*24 hours of EEG data to have a balanced classification of the two classes each consisting of 20 subjects. Through a LOSOCV process [40], the model was trained on 39 subjects and tested on 24-hour data of a specific subject to assess how each subject’s data can predict an individual’s true class (i.e. 1 for the IVH group, and 0 for the control group). A pooling approach is necessary to compute metrics like Kappa and F1-score, as these metrics cannot be computed when the true class is only 1 or 0, as in a LOSOCV approach.

### Quantifying AM and FM Information in EEG Time Epochs Prior to IVH

Analysis of small sample sizes can be biased by subject-to-subject variability. Magri et al [41] proposed the use of mutual information using Shannon’s [42] formula to account for the subject-to-subject response variability (i.e. noise entropy) by subtracting it from the total response entropy, to reliably assess feature contributions despite the variability that may arise due to a limited sample size (see Supplementary section for more details).

If we consider IVH occurrence as a stimulus variable, then there are 20 subjects under the influence of the same stimulus (i.e. 24 epochs of 1h leading up to IVH detection). For example, the 4^th^ epoch of the 24-epoch vector (for all subjects) can be considered as the epoch which is 20 hours before IVH occurrence. Then, we can compute the probability distribution for each of these epochs (based on the feature values) and compute mutual information through the 24 timelines to the IVH occurrence. Bootstrapping the extracted features was applied across the 1-hour EEG epochs (by a factor of 20) and discretized the feature values into 30 levels. These levels were determined heuristically, ensuring the least quantization level while maintaining process repeatability [41–43].

We evaluated the mutual information of all 34 features with the 24 segment stimuli (epochs leading up to IVH occurrence) using I (pre-IVH dynamics; features) ≜ Information (stimulus; response). Subsequently, we determined the joint mutual information of the AM-FM features and stimuli and calculated the corresponding synergy/redundancy. This involved quantifying the increased or decreased amount of information that joint features generated compared to the summation of the individual information of the AM-FM features.

## Results

Table 1 shows that cases were successfully matched and there is no obvious bias between the cases and controls.

### Inter-Subject Analysis

In Figure 2, the classification accuracy and (Cohen’s) Kappa values are presented for 1-hour epochs, encompassing 24 epochs from a cohort of 40 subjects. Among the classifiers evaluated, the AdaBoost method exhibited the highest accuracy and Kappa values. AdaBoost outperformed KNN by over 7% in terms of Kappa values. This outcome underscores the efficacy of considering each epoch as an independent and identically distributed time window, enabling effective discrimination between EEG tracings of infants who develop IVH and who do not.

**Figure 2.**
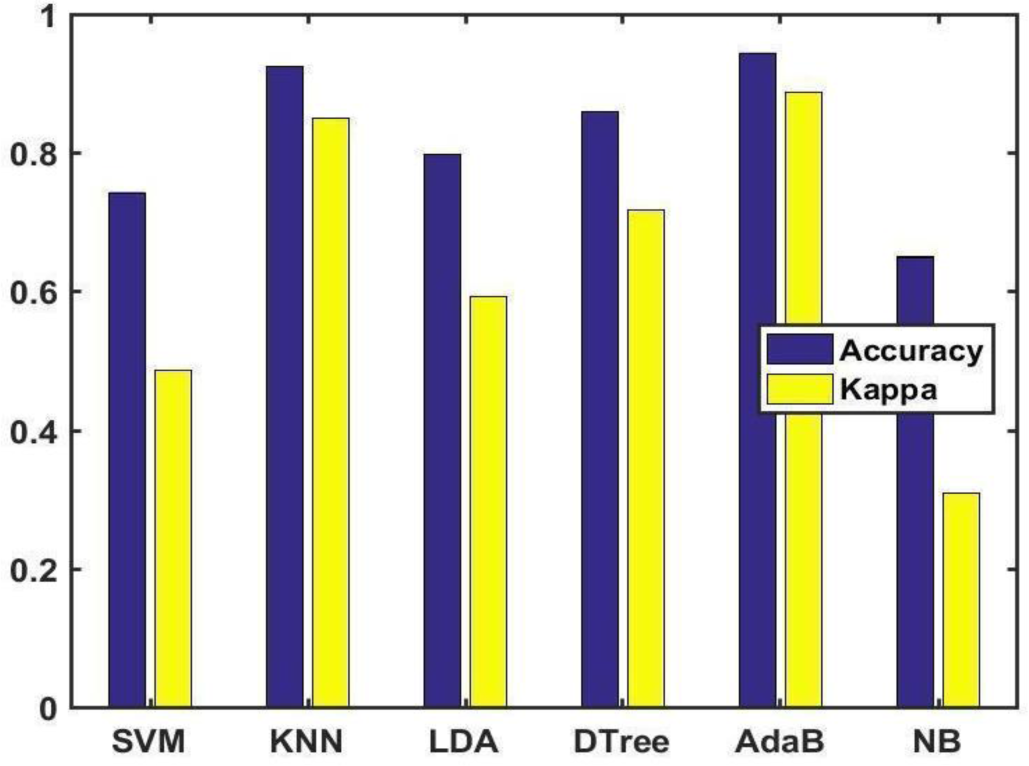
Classification Accuracy and Kappa for 6 classifiers when all the data was pooled. Each EEG was split into 1-h epochs. SVM: support vector machine; 2) KNN: K-nearest neighbours; 3) LDA: linear discriminant analysis; 4) DTree; decision tree learning; 5) AdaB: AdaBoost, and 6) NB: Naïve Bayes.

Figure 3 visually demonstrates the discriminative power of individual features with respect to IVH and control groups. (Table 3) highlights key performance indexes for the AdaBoost classifier, displaying its efficacy in classification tasks. The AdaBoost classifier exceeded the 85% threshold for accuracy. High values of Cohen’s Kappa and F1-Score ensure that if cross-validation folds are chosen not balanced (between the control and IVH group), the weighted accuracy is still high.

**Figure 3.**
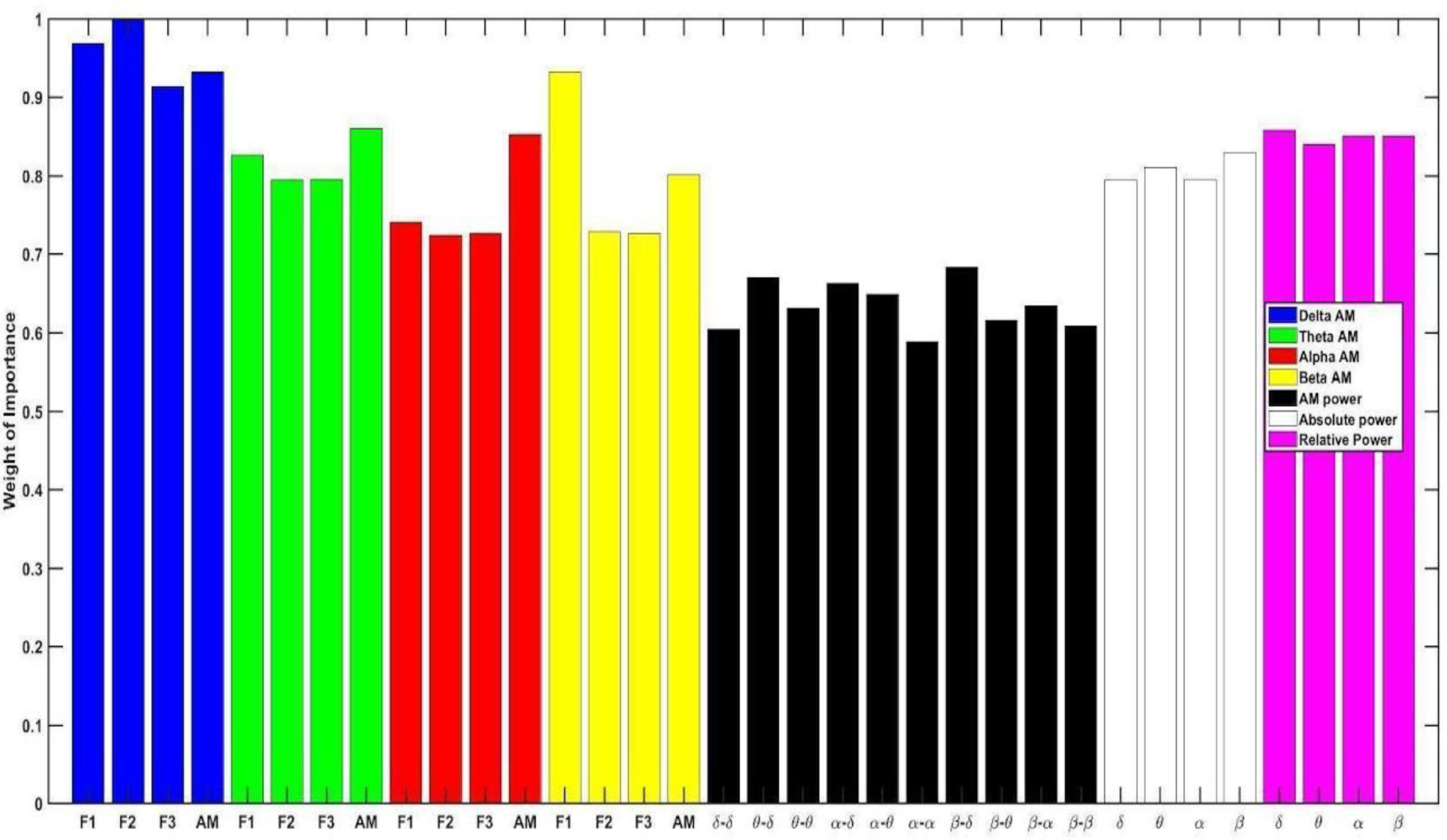
Importance of any single feature for AdaBoost Classification when all the data was pooled. Each EEG was split into 1-h epochs. F1=FM feature. F2=Slow FM feature. F3= Phase Slip FM feature

**Table 3.**
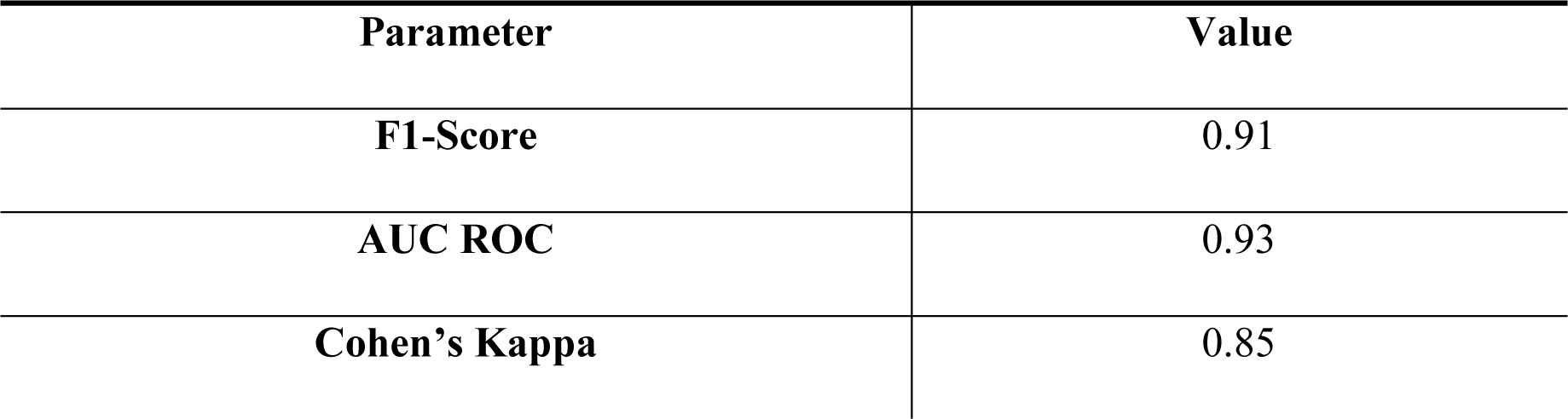
Performance indicators of adaptive boosting algorithm.

### Intra-Subject Analysis

Figure 4 illustrates the accuracy of results obtained for the LOSOCV scenario. The box plot visually represents the range of accuracies achieved for all the epochs across the 20 subjects, within the IVH and control groups. Most subjects (17/20 for the control and 19/20 for the IVH group) achieved an accuracy level exceeding 70% in the LOSOCV case. This observation suggests that the selected features exhibit sensitivity to dynamic changes preceding the occurrence of IVH, leading to favourable true positive classifications. Detailed prediction for each subject and trend of classification (true or false) for the IVH group is depicted in Supplementary figures S2 and S3.

**Figure 4.**
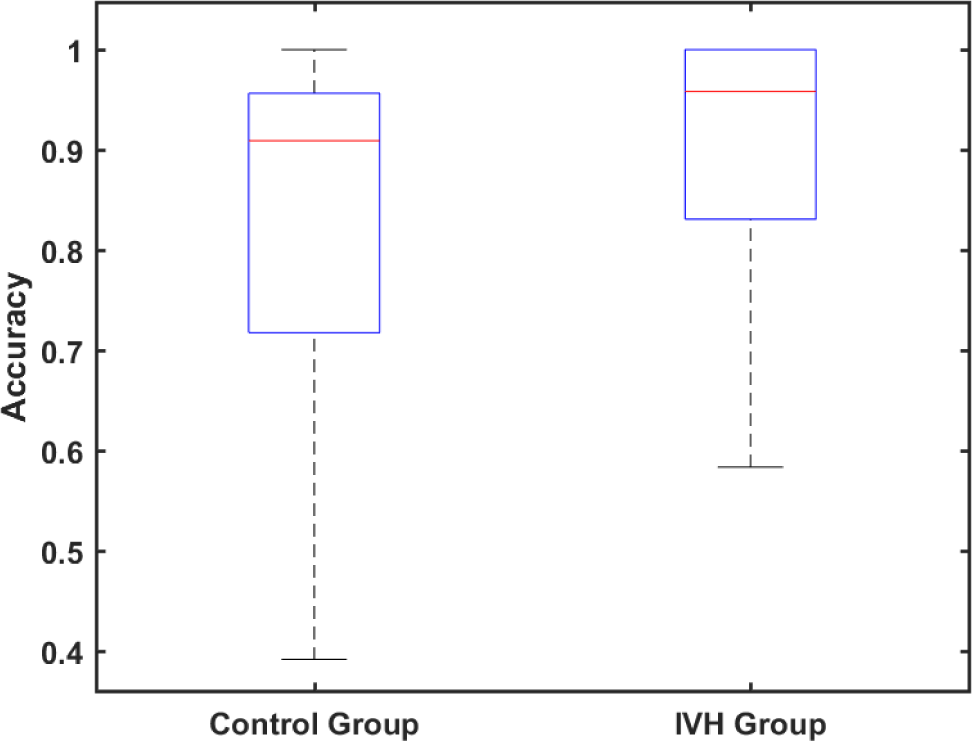
Box plot for accuracy of prediction for each 24-hour dataset of IVH and control group subjects by LOSOCV method.

### Information and Redundancy Analysis

Figure 5 presents the information computed through the 24 epochs (across 20 IVH group subjects) for each of the 34 features individually. Our findings indicate that the AM-FM features in the Delta band remain highly informative for EEG epochs preceding the occurrence of IVH. This observation highlights the robustness of our classification results, as the AM-FM features continue to exhibit significant discriminative power. Figure 6 graphically illustrates the computation of joint information and redundancy. By analysing the joint information and redundancy, we gain insights into the interdependencies and relevance of the EEG epochs and features in our classification task.

**Figure 5.**
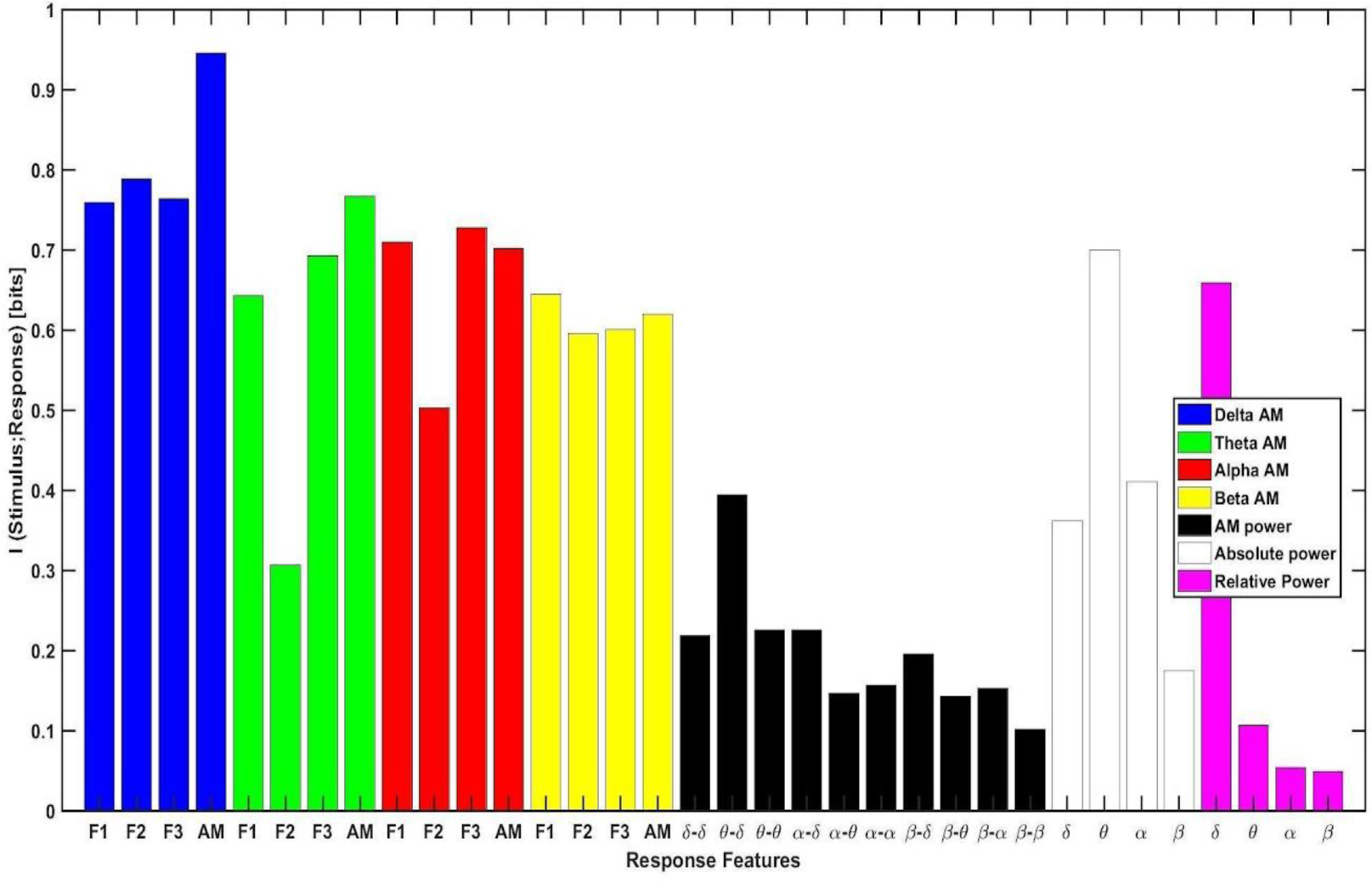
Information computation of 20 subjects’ EEG features prior to IVH occurrence based on each single feature. Stimulus refers to EEG epochs approaching IVH and response is related to the values of each feature. Each EEG was split into 1-hour epochs. F1=FM feature. F2=Slow FM feature. F3= Phase Slip FM feature.

**Figure 6.**
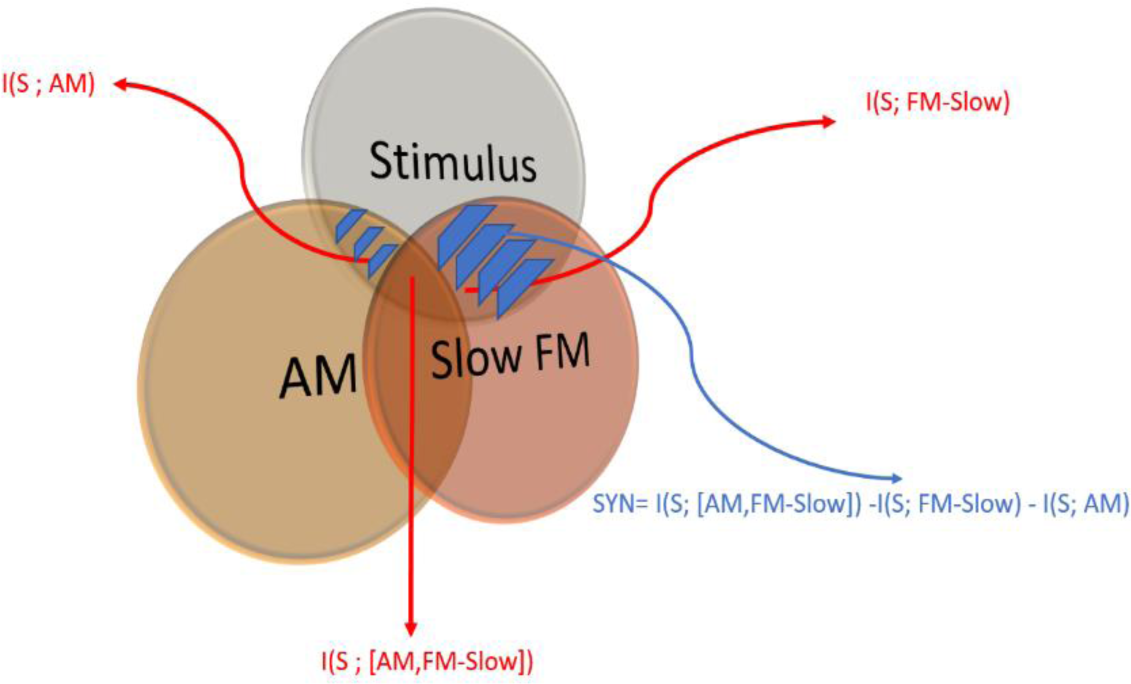
Venn Diagram for general graphical representation of SYN computation for the two dominant features of Slow-FM and AM. S stands for stimulus (upcoming IVH occurrence).

In Table 4, the SYN (synergy if positive and redundancy if negative) values for different EEG epoch lengths are presented. One-hour epochs leads to the lowest redundancy, as indicated by an absolute magnitude of -0.24 bits and a ratio relative to the summation of information of delta AM-FM features (<8%).

**Table 4.**
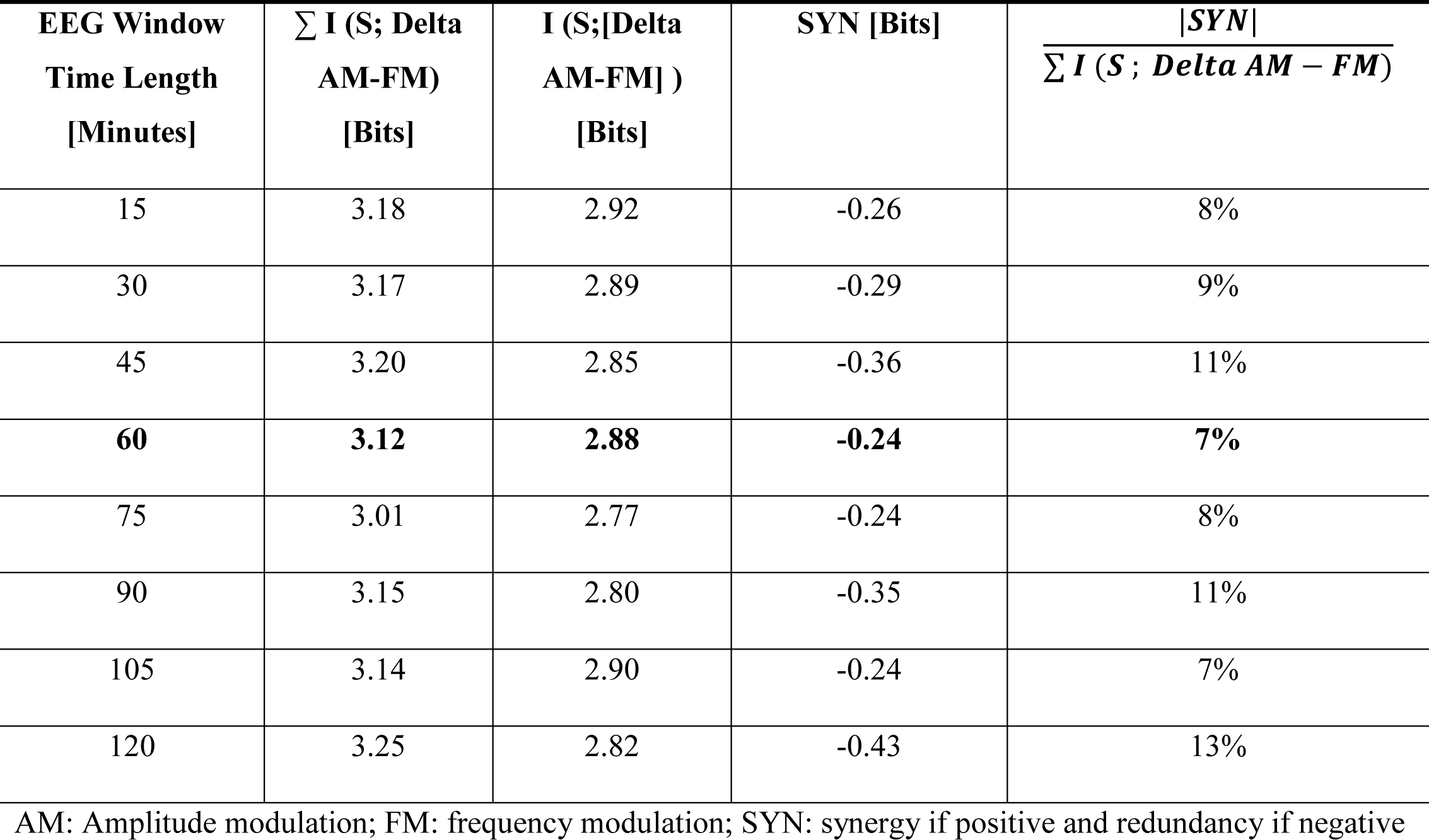
SYN Calculation for different EEG time epochs and the 4 Delta band AM-FM features (FM, Slow FM, Phase slip, and AM)

## Discussion

We introduced amplitude and frequency modulation as a novel method for predicting the occurrence of IVH in preterm infants by analysing a single-channel EEG over a 24-hour period prior to the detection of IVH. To the best of our knowledge, this is the first study to utilize a 24-hour timeline prior to IVH prediction. The 24-hour timeline was demarcated by a clear negative and clear positive cUS, which is difficult to achieve as most clinics do not perform a routine cUS at admission and subsequently daily thereafter. The classification accuracy exceeded 90%. Furthermore, our model achieved an 80% performance in 18 out of 20 subjects within the IVH group, with a maximum of 4 false negatives observed across 24 epochs. For the control group, 17 out of 20 subjects achieved at least 70% accuracy, with a maximum of 6 false alarms out of 24 epochs.

The results demonstrate that slow FM (Delta band) EEG features are useful in accurately predicting the occurrence of IVH in preterm infants. In addition, AM (Delta band) is the most informative feature (0.2 bit more informative than the most informative power band features). Our study not only complements the findings of Iyer et al [23], but also provides new insights into predicting IVH in preterm infants. While Iyer et al [23] focused on burst shape analysis for detecting IVH, our approach utilizes AM-FM features extracted from single-channel EEG signals, providing a different perspective. We demonstrate the additional use of slow FM features in the Delta band for accurately predicting IVH.

Our findings suggest that infants predominantly exhibit informative brain oscillations characterized by lower frequency components, in comparison to adults. [44, 45] Interestingly, our findings indicate that the Delta frequency band [0.1-3.5 Hz] is the primary frequency range for predicting IVH using AM-FM features in preterm infants. In contrast, previous studies in adults have demonstrated significant results in the Beta band when examining AM-FM features [29, 30]. Our results highlight the importance of considering the unique characteristics of neonatal brain oscillations, which differ from those of adults [32, 46].

A possible explanation for our findings is that the sonographer performing the admission cUS (and the reviewers) may have missed an already present but still small IVH [6]. To minimize this bias, original images were re-evaluated by two experienced neuro-neonatologists (JD, MLT) Although unlikely, AM-FM features changes could have reflected the primary IVH, as previously observed in studies reporting EEG changes due to IVH [47–49]. Nonetheless, the novelty of using AM-FM features is their ability to detect EEG changes in the initial stages of IVH, or even prior to IVH.

This study has potentially important implications for clinical practice and future research. Currently, several approaches have been suggested for reducing the incidence of IVH in preterm infants, through the implementation of nursing intervention bundles during the first 72 postnatal hours. The effectiveness of such interventions was demonstrated in a previous study by de Bijl-Marcus et al [50], which showed a significant reduction in both low-grade and severe haemorrhages in the intervention group. Identifying those infants who are at the highest risk could further increase personalized neuroprotective strategies, and hopefully prevent IVH or reduce the risk of complications of IVH (e.g. venous infarctions, PHVD) [51]. Although our study results are very promising, implementation might be challenging due to the tendency of reduced aEEG monitoring in infants born extremely preterm, as electrode placement is considered to be a burden.

Several limitations need to be discussed. First, we encountered difficulties in selecting preterm infants with IVH that had a prior negative cUS, while having sufficiently high-quality EEG data available prior to IVH detection. Therefore, we present a pilot study of 20 infants with IVH that was not present directly after birth, and who were successfully matched to control infants who did not develop IVH. Second, figure 4 indicates that the accuracy of the algorithm was less than 70% in two cases. Nevertheless, all 24 epochs were classified and estimated without error in 12 cases. A larger number of subjects in future studies may help to address this limitation and enable (external) validation and generalizability of the findings. Finally, a retrospective study using cUS as an outcome measure for detecting IVH may include inconsistent timing, potential variability in image interpretation and reporting, and potential selection bias due to the exclusion of infants who did not receive their routine ultrasound scans. As discussed before, obtaining a dataset as presented here is challenging, and the presented data is rather unique.

Considering the future implications, our study introduces the promising and innovative approach of AM and FM features for dynamically predicting IVH in preterm infants. This method holds the potential to enhance individualized care and might improve patient outcomes, which would be a significant advancement in the field. By continuously tracking the time course of pre-and post-IVH occurrence, we can gain a more thorough understanding of the dynamic shifts that occur within the brain and extract richer insights into the pathophysiology of this condition. Furthermore, the use of more complex brain signals, such as AM-FM features, opens new possibilities for predicting IVH and understanding the mechanisms of birth maturity reflected in brain dynamics. However, there is still much to investigate regarding the potential of neural activity-derived biomarkers and their association with brain injury in preterm infants. Therefore, our study provides a foundation for future research in this area.

## Conclusion

Our pilot study provides strong evidence supporting the use of quantitative EEG analysis by means of AM-FM modulation for predicting the development of IVH in preterm infants within the 24-hour window prior to its detection. This robust evidence underscores the potential of our approach for early identification of IVH and intervention, offering promising prospects for enhancing the management and care of preterm infants at risk of IVH.

## Data Availability

Anonymized data and codes are available upon request to the corresponding author.

## Abbreviations

IVH: Intraventricular Haemorrhage
EEG: Electroencephalography
AM: Amplitude Modulation
FM: Frequency Modulation
LOSOCV: Leave-One-Subject-Out Cross Validation
PHVD: Post Haemorrhagic Ventricle Dilation
SYN: synergy
CCS: corticosteroids
EOS: microbiologically proven sepsis <48h of age
HELLP: hemolysis elevated liver enzymes and low platelets syndrome
HSPDA: hemodynamically significant patent ductus arteriosus
IQR: interquartile range;
LOS: microbiologically proven sepsis >48h of age
NEC: necrotizing enterocolitis
PE: preeclampsia
PVHI: periventricular hemorrhagic infarction
SD: standard deviation
SGA: small for gestational age

## Summary

This study aimed to investigate the potential of amplitude and frequency modulation (AM-FM) features in quantitative EEG analysis for predicting intraventricular haemorrhage (IVH) in preterm infants. IVH is a complication of preterm birth that can lead to severe disabling neurological sequalae, and its early detection is crucial for timely intervention to minimize brain injury and improve patient outcomes.

The study was conducted as a case-control analysis on preterm infants with a gestational age (GA) less than 30 weeks. All infants underwent routine amplitude-integrated EEG monitoring for at least 3 days after birth. The cases were included when there was an IVH visible on cranial ultrasound and when they had an ultrasound 24h before that did not demonstrate an IVH. The cases were matched with controls, without IVH, based on sex, GA, and birth weight. The EEG data was analysed in 1h-epochs. Six machine learning algorithms were evaluated to develop a predictive model using AM-FM features extracted from single-channel EEG data. Algorithms that were evaluated: 1) support vector machine; 2) K-nearest neighbours; 3) linear discriminant analysis; 4) decision tree learning; 5) AdaBoost, and 6) Naïve Bayes. A 10-fold cross validation and leave-one-subject-out-cross-validation was performed to validate the model. Optimal EEG epoch length was evaluated by means of a “synergy if positive and redundancy if negative” analysis.

The findings demonstrated that AM-FM modulation features of EEG data 24 hours before IVH detection were predictive of IVH occurrence. The predictive model using adaptive boosting achieved a remarkable area under the receiver operating characteristic curve of 93%, an accuracy of 91%, and a Kappa value of 0.85. Notably, the slow varying instantaneous frequency and amplitude features in the Delta frequency band showed the most significant mutual information and little redundancy, making them valuable biomarkers for IVH prediction.

The study’s results indicate that AM-FM modulation of EEG signals provides valuable insights into neural activity and dynamics, making it a potential tool for predicting IVH in preterm infants. The utilization of AM-FM features in EEG analysis yields new opportunities for early identification and intervention in at-risk infants, enhancing their management and care. Moreover, this approach offers the possibility of personalized information for clinical decision-making, optimizing the prediction accuracy for each individual subject.

Overall, this pilot study contributes to the understanding of using AM-FM features in EEG analysis as a promising method for predicting IVH in preterm infants. The evidence gathered supports the potential benefits of proactive IVH prediction, allowing for timely interventions to prevent or minimize brain injury in this vulnerable population. Further research and validation on a larger scale are warranted to solidify the use of AM-FM modulation as a valuable tool in neonatal care, providing clinicians with critical insights to improve long-term neurological outcomes for preterm infants.

