## Supplementary for "Amplitude and Frequency Modulation of EEG Predicts Intraventricular Haemorrhage in Preterm Infants"

### Amplitude- and frequency modulation features

The amplitude (AM) and frequency modulation (FM) features (first 4 rows of features in Table 2) are based on Averna et al. [29]. The filtered EEG signal assumed as x(t) (which has duration of 1-h for each epoch), was transformed using the Hilbert transform to obtain its analytic signal z(t), from which the instantaneous phase θ(t), amplitude A(t), and frequency f(t) were calculated:

$$z(t) = x(t) + j^*H[x(t)] \quad (1)$$

$$\theta(t) = Arc\tan[H[x(t)]/x(t)] \quad (2)$$

$$A(t) = |z(t)| = \mathrm{sqrt}\left(\mathrm{Re}\big(z(t)\big)^2 + \mathrm{Im}\big(z(t)\big)^2\right) \quad (3)$$

The AM and FM were quantified using the log-transformed variance of the instantaneous amplitude and the variance of the instantaneous frequency, respectively. In the presence of phase slips, the instantaneous frequency was decomposed into slow FM and phase-slips FM using cubic interpolation.

$$AM = \log(\mathrm{Var}[A(t)]) \quad (4)$$

$$FM = Var[f(t)] \quad (5)$$

The AM power features (5-th row of features in Table 2) are based on Fraga et al. [30]. First, the full-band EEG signal was decomposed into four sub-bands: the delta (0.5-4 Hz), theta (4-8 Hz), alpha (8-12 Hz), and beta (12-30 Hz). The temporal envelope of each sub-band EEG signal was then computed by means of a Hilbert transform Let x_i(t) denote the temporal envelope of the $i^{th}$ sub-band signal, where i=1,2,3,4.

To quantify the temporal dynamics of the sub-band envelopes, we perform a second frequency decomposition into five so-called modulation bands using second-order bandpass modulation filters (with quality factor Q~2). Let y_i(t) denote the temporal envelope of the $i^{th}$ modulation band signal, where i=1,2,3,4. The resulting frequency-frequency signal representation conveys rate-of-change information of each of the sub-band envelopes.

The following cross-frequency amplitude modulation interactions were computed:

- E(delta; m-delta)
- E(theta; m-delta, m-theta)
- E(alpha; m-delta, m-theta)
- E(beta; m-delta, m-theta, m-alpha, m-beta)

In our experiments, these 10-percentage cross-frequency modulation parameters are computed for each of the EEG epochs. For a schematic of AM power calculation refer to Fraga et al. [30]. We did not use the Gamma band because it was not significant in neonates' neural oscillations.

Fig. S1 depicts one example of a 40 second EEG signal and the corresponding AM-FM dynamics of it. It is obvious that there are fast and slow dominant varying instantaneous frequency components (in Delta band) which are not easily recognized by the raw signal or bi its bandpass filtered outcome.

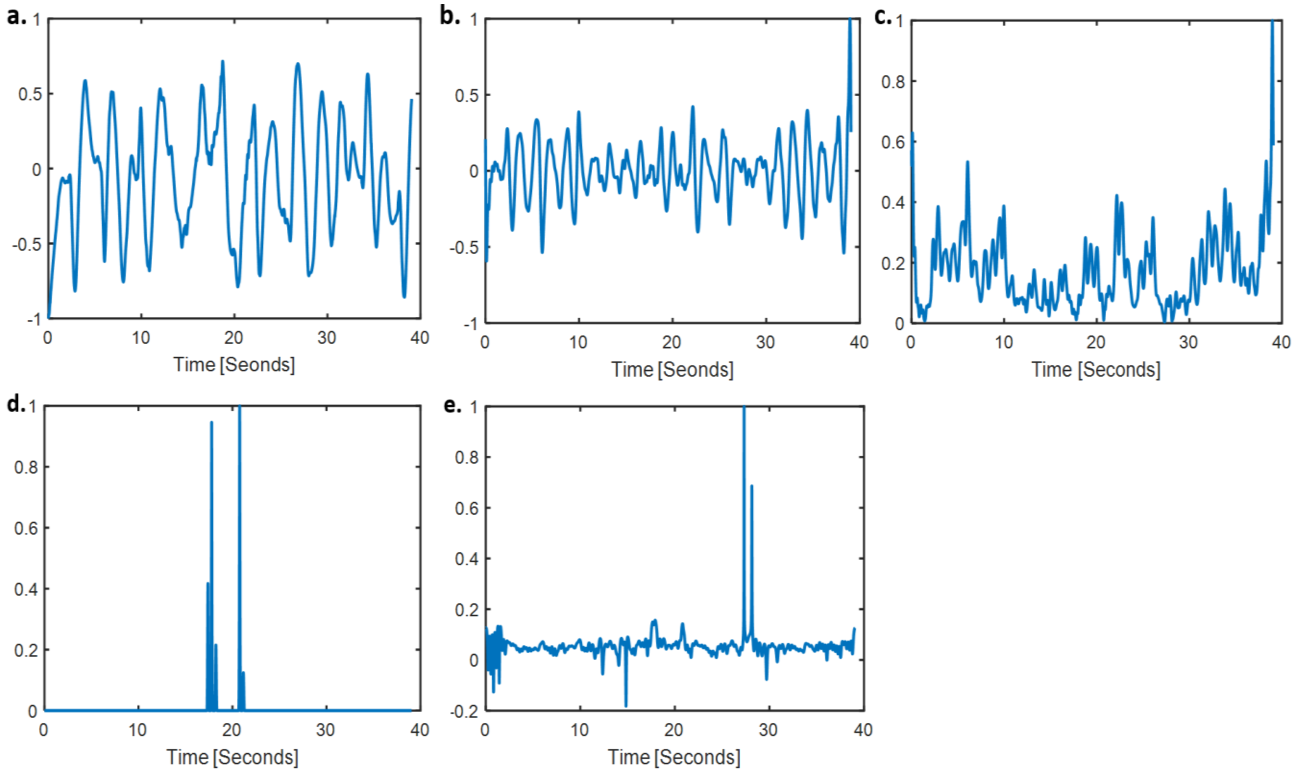


Figure S1. Example of an AM-FM signal extraction from EEG signal in Delta band (normalized values). a) EEG signal. b) Delta band filtered EEG signal. c) Instantaneous amplitude. d) (Magnitude of) Phase slip of FM. e) Instantaneous frequency.

### Hyper-parameter optimization

Table S1 is the results of optimized parameters for 6 classifiers. These are the parameters (found by Gridsearch package) to work based for classifying between IVH and control groups. The fine-tuned parameters (out of Gridsearch) were specifically tailored for the classification task of distinguishing between intraventricular haemorrhage (IVH) and control groups within the study.

Table S1. Best learning parameters for each classification method obtained by Grid Search

| Model | Best Parameters |
|---|---|
| SVM | {'Kernel': Linear}; {'Box constraints'=0.0016}; {'Solver': SMO}; {'Kernel Function'=Polynomial}; {'Polynomial Order': 3} |
| Naive Bayes | {'Kernel': Normal – 'Width'=187.27} |
| Adaptive Boosting | {'Number of learning cycles'=437}one; {'Learning rate'= 0.77}; {'Minimum Leaf Size'=4}; {'Max Splits'=58} |
| Discriminant | {'Discriminant type': Pseudo linear}; {'Gamma'= .65}; {'Delta'=.0001} |
| K-nearest neighbours | {'Distance': Minkowski}; {'Distance weight': Inverse}; {'Number of Neighbours'=2}; {'Exponent':0.67} |
| Decision Tree | {'Split criterion': Deviance}; {'Minimum Leaf'=4}; {'Max Splits'=4} |

Among the 6 classifiers, as notable in the next section, the adaptive boosting algorithm performed best, demonstrating a high learning rate of 0.77. A higher learning rate in adaptive boosting signifies that each weak learner exerts a more substantial influence on the final prediction, potentially resulting in faster convergence. However, it is important to consider the associated trade-off, as this elevated learning rate also increases the risk of overfitting. To further investigate this possibility, the subsequent section includes an analysis of Kappa values, which will provide valuable insights into the classifier's performance.

### Mutual information and synergy

Mutual information is a measure of the amount of information shared between two random variables, in this case, between EEG epochs and features. It is given by the formula:

$$I(X;Y) = \sum\sum p(x,y) \log_2 [p(x,y) / (p(x)\ p(y))]$$

where X and Y represent the random variables (in this case, EEG epochs and features), and p(x), p(y), and p(x,y) represent their probability distributions. The mutual information I(X;Y) represents the amount of information that knowing one variable (X or Y) provides about the other variable (Y or X).

The mutual information formula includes two terms: the response entropy and the noise entropy. The response entropy captures the overall variability of the response and is calculated based on the probability of observing the response r for any stimulus. The noise entropy quantifies the response variability that is specifically due to "noise," or trial-to-trial differences in the response to the same stimulus. To calculate the noise entropy, we use the conditional entropy formula, which incorporates the probability of observing a response r given a stimulus s and the probability of presenting the stimulus s. The probability of presenting the stimulus s is determined by the number of trials available for that stimulus divided by the total number of trials for all stimuli.

The mutual information measurement provides an estimation of how much of the information capacity conveyed by stimulus-induced differences in neural activity is resistant to the trial-by-trial variability of the response. Alternatively, it describes the extent to which the uncertainty about the stimulus can be reduced by observing a single trial of the neural response.

Redundancy is a measure of the amount of overlap between two sources of information. It is related to the concept of synergy, which refers to the amount of information that is shared between two sources beyond what would be expected based on their individual contributions. In this case, we can compute redundancy between EEG epochs and features using the formula:

$$R(X;Y) = I(X;Y) - H(X|Y)$$

where H(X|Y) is the conditional entropy of X given Y and represents the amount of uncertainty in X that remains after Y is known. The quantity R(X;Y) represents the amount of information that is shared between X and Y, beyond what would be expected based on their individual contributions. If synergy is negative, then there is redundancy between X and Y, meaning that knowing one variable provides some information about the other variable. If R(X;Y) is negative, then there is redundancy between X and Y, meaning that knowing both variables provides more information than would be expected based on their individual contributions.

### Detailed classification trend among IVH group

To see how classification works for each 1-hour epoch, Fig A3, shows the scatter plot of classification situation through time.

The average over 20 subjects for each epoch could potentially be a measure for prediction accuracy through time. However, because measurement with CUS is not continuous, it is not able to synchronize timeline epochs before IVH occurrence among the IVH group subjects. We averaged the classification accuracy among 4 hours windows (4*20 values averaged) for a better trend analysis. It seems like the error is low at first and at time approaching IVH. In middle the error is still low but a little higher than boarders of 24 hours timeline.

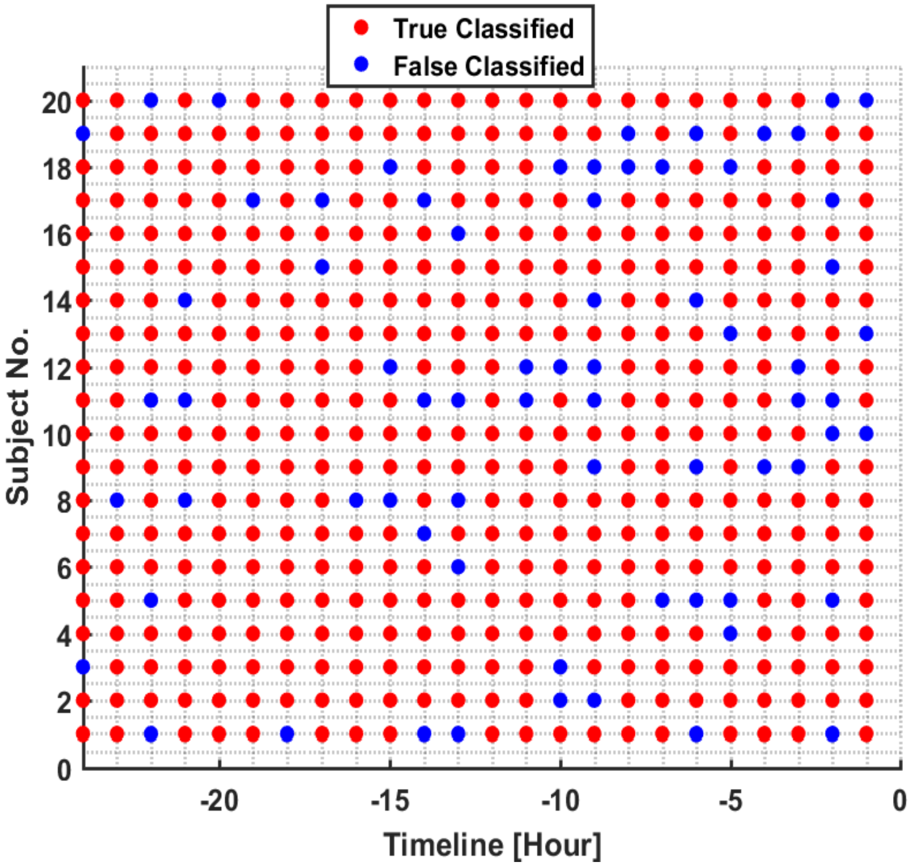


Figure S2. Scatter plot of classification through time toward IVH occurrence. Timeline -X, means X hours before IVH occurrence.

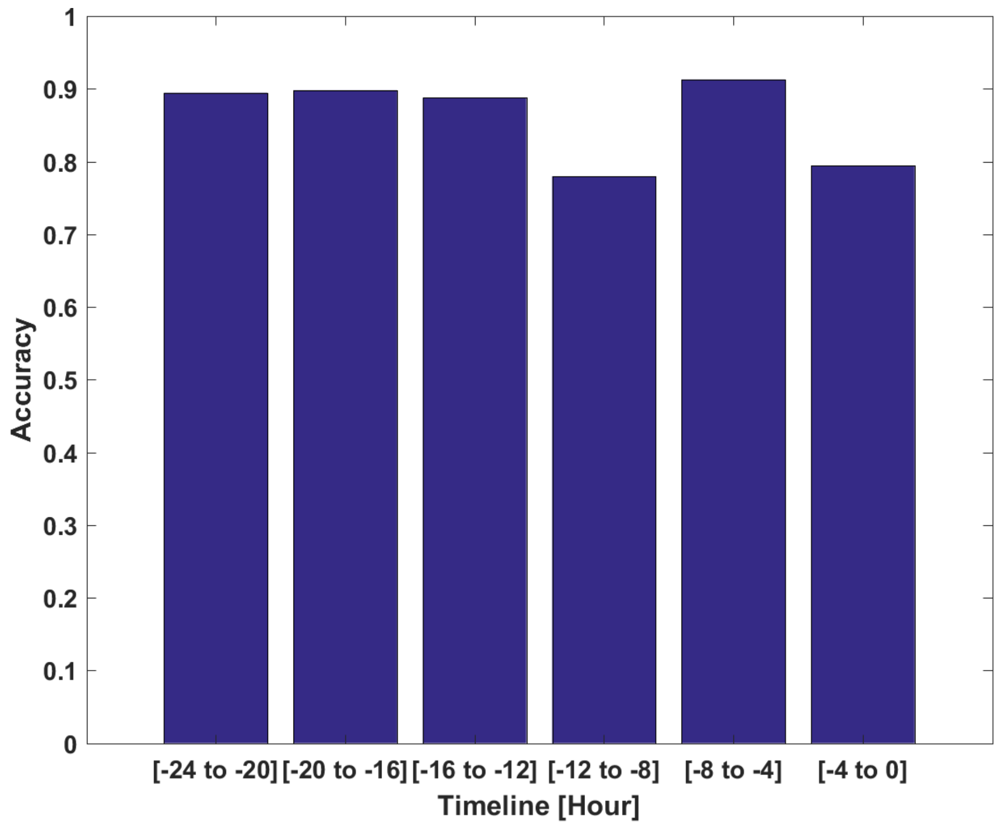


Figure S3. bar plot of average classification accuracy through time toward IVH occurrence. Timeline -X, means X hours before IVH occurrence.
